# Personalizing Mobile Applications for Health Behavioral Change according to personality: cross-sectional validation of a Preference matrix

**DOI:** 10.1101/2025.05.07.25327187

**Authors:** Laëtitia Gosetto, Gilles Falquet, Fréderic Ehrler

**Author notes:** **Corresponding author:** (LG).

## Abstract

mHealth apps are increasingly popular, offering tools like health tracking and personalized reminders to support these behaviors. Personalized messaging, tailored to the user’s profile, has been shown to improve engagement and retention around health topics. Research links personality traits (based on the Big Five model) with preferred app mechanisms, leading to a preference matrix for personalizing health apps. This preference matrix includes 15 mechanisms, categorized by the Behavior Change Technique and gamification elements, guiding developers to optimize app engagement based on user profiles. This study aims to validate this preference matrix by assessing whether the associations between mechanisms and Big Five personality profiles proposed in the preference matrix align with the preferences of our population in an experimental context. This study employs a cross-sectional design. Participants completed an online survey, which collected data on demographic information, mobile health app usage, and personality. The average age of the 214 respondents (118 women, 89 men, 5 others), was 29.42. Logistic regression and logistic ordinal regression analyses, adjusted using the Bonferroni correction, assessed the influence of personality traits on mechanism preferences and motivation levels was statistically significant for three mechanisms. Conscientiousness significantly increased the likelihood of selection for collection (OR = 1.87). For competition, both conscientiousness (OR = 3.22) and altruism (OR = 1.93) emerged as strong predictors. For rewards, conscientiousness (OR = 1.97) and neuroticism (OR = 2.36) also showed a strong predictive value. The study found that four mechanisms—self-monitoring, progression, challenge, and quest—were favored by over half of the participants, suggesting these should be standard features in mHealth applications. Conscientious participants showed a preference for the collection mechanism, while both conscientious and altruistic individuals were drawn to competition. Neurotic and conscientious individuals preferred the reward mechanism. Conscientiousness consistently predicted preferences for all three gamification elements, highlighting its role in influencing engagement with mHealth features.

**Author summary:** The increasing emphasis on fostering healthy behaviors and mitigating health-related risk factors—such as physical inactivity, a major contributor to mortality—has driven the rapid expansion of mobile health (mHealth) applications. These digital tools are designed to support behavior modification by incorporating functionalities like reminders, goal tracking, and individualized intervention strategies, which have been associated with improved user engagement and sustained adherence. Central to the efficacy of these interventions is personalization, wherein adaptive content is aligned with the unique attributes and preferences of individual users. Empirical findings indicate that customized messages and feedback outperform standardized approaches, particularly in the promotion of physical activity and weight management. Moreover, the integration of gamification elements is a common strategy employed to enhance motivation and user interaction, with preliminary evidence supporting its positive influence. In this study, we seek to enhance the impact of digital health interventions by systematically mapping game mechanics, behavioral change strategies, and application features to user profiles characterized by the Big Five personality dimensions.

## Introduction

Many studies have demonstrated the role of healthy lifestyle behaviors adoption to increase life expectancy. Specifically, the adoption of a healthy diet, maintenance of a healthy weight, cessation of smoking, consumption of alcohol in moderation, and regular exercise are associated with a reduction in mortality. The researchers discovered that the adoption of each healthy lifestyle behavior contributes to an enhanced quality of life and longevity [1].

The number of health apps designed to assist individuals in adopting healthier behaviors is increasing annually, with new apps reaching the market every year. In 2018, there were over 35,000 health apps available for download [2]. The advent of smartphone apps has created novel avenues for individuals to adopt health-related behaviors. These apps offer immediate access to health-related information, medication reminders, and tools for monitoring progress, which can collectively fostering healthier lifestyles [3].

Several scales have been developed to assess the quality of mHealth apps. For example, the Mobile App Rating Scale (MARS) [4] and App Behavior Change Scale [5]. One of the most salient features of these scales is the inclusion of personalization as a quality factor. Indeed, personalization represents a crucial consideration when developing an app designed to facilitate behavioral change. For example, one study has demonstrated that messages tailored to the user’s specific characteristics are more likely to be read, recalled, and retained in memory, and are perceived as more personally relevant than untailored messages. Additionally, tailored messages tend to garner more attention, elicit more discussion, and be a topic of interest among others, compared to untailored messages [6].

### The preference matrix

Personalization of mobile app can be done according to the user’s profile. In previous articles [7], we identified from the literature preference relations between personality traits and mechanisms (components can be incorporated within a mHealth application). These findings related to the development of a preference matrix indicating the type of mechanisms preferred according to a user profile. The preference matrix can be used as a framework for the customization of mobile applications with the objective of encouraging behavioral change. When designing a mobile application aimed at facilitating behavior change for health-related purposes, designers can utilize our preference matrix to identify the mechanisms that are most efficient to achieve the desired outcomes, given the profile of the intended users. For instance, if an individual is classified as extroverted according to the Big Five personality traits, it would be relevant to prioritize mechanisms that facilitate social comparison and collaboration between users [8].

The preference matrix includes 15 mechanisms linked to Big five personality profile, which are presented in detail in Table 3. Regarding the user profile, we utilized one of the most prevalent classification systems for individuals, namely the Big Five personality profiles [8–16].

**Table 1.** Definition of the Big Five factors personality.

| Traits | The five traits represent the tendency to [17] ... |
| --- | --- |
| Neuroticism | Worried, nervous, emotional, anxious, maladjusted, hypochondriac |
| Extraversion | Sociable, active, talkative, open to others, optimistic, fun-loving, affectionate |
| Openness | Curious, eclectic, creative, original, imaginative, non-conformist |
| Altruism | Compassionate, easy-going, trusting, helpful, indulgent, credulous, honest |
| Conscientiousness | Organized, reliable, hard-working, disciplined, punctual, meticulous, careful, ambitious, persevering |

**Table 2.** List of mechanisms with their definition.

| Mechanism | Definition |
| --- | --- |
| **BCT mechanisms** |  |
| Prompt and cues | Usually a message delivered to the user to prompt or recall a behavior at a specified time, with the app or user defining when the message should be sent [21] |
| Demonstration of the behavior | Enables users to observe the cause-and-effect linkage of their behavior, such as seeing a simulation of their bodies after a diet [22]. |
| Self-monitoring | Users can track their behaviors, providing information on both past and current activities [22] |
| punishment | Virtually penalizes the user for not performing the desired behavior or reaching their goal [10] |
| Social comparison | An individual's perceptions of the prevailing beliefs and behaviors within a social group. |
| social support | Enables communication between users, such as through chat or sharing activities with other users [23]. |
| **Game elements** |  |
| Progression | Users can track their progression with steps through the system's purpose over time, visualized with mechanisms like stars or flags along a path [22]. |
| Competition | Users can compete to accomplish the desired behavior [22]. |
| Cooperation | Users collaborate to achieve a shared objective [22]. |
| Collection | Allows users to gather virtual objects. |
| Rewards | Virtual rewards offered to users for engaging in the target behavior [22]. |
| Quest | Users can enter or define the objectives targeted for the activity they will perform [21]. |
| Challenge | Presents various situations that require effort from the user to be completed [23] (e.g., accomplishing 3 hours of physical activity per week). |
| Avatar | Allows users to share their data in the system without revealing their name [23]. |
| **App mechanism** |  |
| Customization | In contrast to personalization—which involves adjusting automatically the system to the user—customization refers to the user's ability to modify the content or functionalities of the mobile application according to their own preferences [22]. This approach enables users to actively tailor the system based on users' choices. |

**Table 3.** Relation between big five traits and mechanisms.

|  |  |  | BCT mechanisms |  |  |  |  |  | Game elements |  |  |  |  |  |  |  | App mecha<br>nism |
| --- | --- | --- | --- | --- | --- | --- | --- | --- | --- | --- | --- | --- | --- | --- | --- | --- | --- |
|  |  |  | Prompts and cues | Demonstration of the behavior | Self monitoring | Punishment | Social comparaiso | Social support | progression | Competition | Cooperation | Collection | rewards | Quest | Challenge | avatar | customisation |
| Personality profile | Big Five | Openess to experience |  |  | ++ | + | + | ++ |  | +++ - | + - | ++ | ++ - |  |  |  | ++<br>+ |
|  |  | Agreeableness | + | + | +++ | ++ | ++ - | +++ | ++ | +++ - | +++ | ++ | +++<br>+ |  | ++ |  | +++ |
|  |  | Conscientiousness |  | + | + - |  |  |  | + | ++ | - - |  | ++ | + | + |  | - |
|  |  | Extraversion | + | + | ++ | + | +++ | ++ | +++<br>+ | +++ | ++ | + | +++<br>++ | + | ++ | + | +++<br>- |
|  |  | Neuroticism |  |  | + |  | + | + | + | + - | + | ++ | +++<br>+ |  |  |  | + |

+ 1 article with a preference relation - 1 article with non-preference relation

### Personality measure: the Big-five

One of the most widely used instruments for the assessment of personality is the Big Five Model. The Big Five model characterizes an individual’s personality based on five distinct traits, as illustrated in Table 1.

This model has the advantage of being frequently used in academic research. Between 1990 and 1994, the number of publications on this topic was approximately 400, while between 2005 and 2009, this figure rose to approximately 1,600 [18].

The other advantage of this model is its universality. Five-factor model is universal in humans [19]. Indeed, the authors translated their personality questionnaire, the NEO-PI-R, into six languages belonging to the same language family as English (German, Portuguese, Hebrew, Chinese, Korean, and Japanese). The Big Five factors were found to emerge consistently.

In a further study, the researchers investigated whether the Big Five personality dimensions, as measured by the NEO-PI-R, are universal across 50 cultures, including American, European, Arabic, Asian, and African [20]. Accordingly, the NEO-PI-R was translated into the original language of the country in which it was administered. The participants were primarily students from the country in which the test was administered. The factor analysis demonstrated that the American structure of the NEO-PI-R self-report questionnaire (Form S) could be replicated in all cultures, with 94.4% of factors replicated. Furthermore, the structure was recognizable in all cultures studied.

### Selection of the 15 mechanisms

Based on the hypothesis that people with different profile have different preference about components of mHealth applications aiming at facilitating behavioral change, a review of the literature identified 15 mechanisms that have been associated with user profiles. In our context, the term ‘mechanism’ is employed to denote the entirety of components that can be incorporated within an mHealth application with the objective of promoting behavioral change. We classified the mechanisms into two categories, the mechanisms linked to Behavior change techniques and the mechanisms linked to game elements. For further details and definition of mechanisms, refer to Table 2.

### Preferences relations between big-five traits and mechanisms

The preference relations between the mechanisms and the big-five traits found in the literature enable us to build a preference matrix summarized in Tables 3, each cell of the matrix indicating the number of articles of the review containing a preference relation. Most of the preference relations were positive (71%, 53/75), indicating a preference for a specific mechanism within a given profile (represented by a plus sign). Negative relations indicate an aversion for a mechanism (represented by a minus sign in Tables 3) and represent 12% (9/75) of potential relations. It is worth noted that the relation matrix is not yet complete, as 73% (55/75) of potential relations were found in the literature review performed. This study aims to corroborate or enrich this preference matrix with new preference relations the preference matrix displaying relations between mechanisms and personality traits by experimentally.

## Methods

This section comes from our previous published protocol article [24].

### Ethics Approval

The University Ethics Commission has approved this study for ethical research at the University of Geneva (CUREG_2021-04-38).

### Study Design

We performed a cross-sectional study to address our aims. Participants responded to an online questionnaire in accordance with the Checklist for Reporting Results of Internet E-Surveys [25].

### Outcomes

The primary outcome is the preferred mechanisms given the user profile.

### Study Population

The target population for this study are individuals aged 18 and above who understands French. The recruitment process was conducted via social media platforms, specifically Facebook and Twitter, targeting students at the University of Geneva.

### Sample Size calculation

We calculated the required sample size using a multiple regression power analysis in R. The parameters used were the number of predictors u=3, effect size f2=0.07, significance level α=0.05, power = 0.9, and estimated variance = 202.403. These estimates were based on the hypothesis that individuals with higher levels of altruism, as measured by the Big Five personality traits, show a preference for social networks [11,23].

To approximate the variance, we referred to a prior study [26] that assessed Big Five personality traits in relation to user preferences for social network posters. Specifically, we used the variance in Big Five trait ratings for altruistic participants (n = 46) based on their average scores for a poster promoting blood donation, which had a scoring range of 0–100. Using these parameters, we determined that a sample size of 206 would be sufficient for this study.

### Procedure

The participants were requested to complete an online questionnaire, which was developed by the investigators using the Qualtrics software (Qualtrics, Provo, UT) (see Multimedia Appendix 1). First, participants were required to complete a consent form that detailed the purpose of the study, the procedure to be followed, and their right to withdraw from the study at any time. It was compulsory for the participants to confirm that they had read and understood the form and agreed to its terms before to gain access to the questionnaire and for the investigators to utilize the data provided by the participants for the purposes of this study. Subsequently, participants were requested to respond to questions pertaining to their demographic characteristics. In order to access the rest of the survey, participants were required to confirm that they were over the age of 18. Eligible participants proceeded to complete the online questionnaire, which required them to (1) respond to a scale for measuring their personality profile and (2) examine the 15 mechanisms, select their five favorites, and indicate their likelihood of being motivated by a mechanism to get fit on a scale from 0 to 100.

## Measures and Measurement

### Demographic questions

The participants were requested to provide information regarding their gender, age, occupation, and level of education.

### Profile Assessment

To assess the personality of the participants, we relied on the French version of the Big Five Inventory-10 scale (BFI-10-Fr), which was translated from English and validated by Courtois [26]. The internal reliability of the BFI-10 is low, as indicated by Cronbach’s alpha coefficients ranging from 0.37 to 0.83. This is due to the fact that Cronbach’s alpha is not an appropriate method for evaluating scales with a limited number of items [26]. The scale comprises 10 items, with two items pertaining to each of the five Big Five dimensions. Participants were requested to indicate on a 5-point Likert scale whether they approve or disapprove the statements pertaining to themselves. For example, the subject may indicate that they see themselves as reserved or as someone who is easily anxious. Subsequently, the score for each dimension is calculated by adding the scores for the two statements concerning the dimension, with the items reversed.

This scale was selected due to its factorial structure, which is identical to that of the complete version of the BFI-Fr [26]. Consequently, this approach offers the advantage of effectively measuring personality with a limited number of items. Given the number of scales included in our protocol, it was deemed preferable to select the shortest valid versions in order to avoid participant fatigue resulting from a questionnaire that was too lengthy.

### Choice of mechanisms

Presentation and selection of the five favourites mechanisms

A mockup of each 15 mechanisms (see S1 Appendix) was create. A comprehensive presentation of all mechanisms and their definitions can be found in the Multimedia Appendix. In order to limit the potential bias due to design preferences, the visual design of the mockups was intentionally minimalistic. It relied only on a black and white color scheme with icons, in order to ensure neutrality and clarity. (Fig 1). To avoid any primacy or recency effects, the 15 mockups were randomly presented to the participants. During the study, participants were required to select the five mechanisms they considered the most motivating based on its mockup accompanied by a brief textual description.

**Fig 1.**
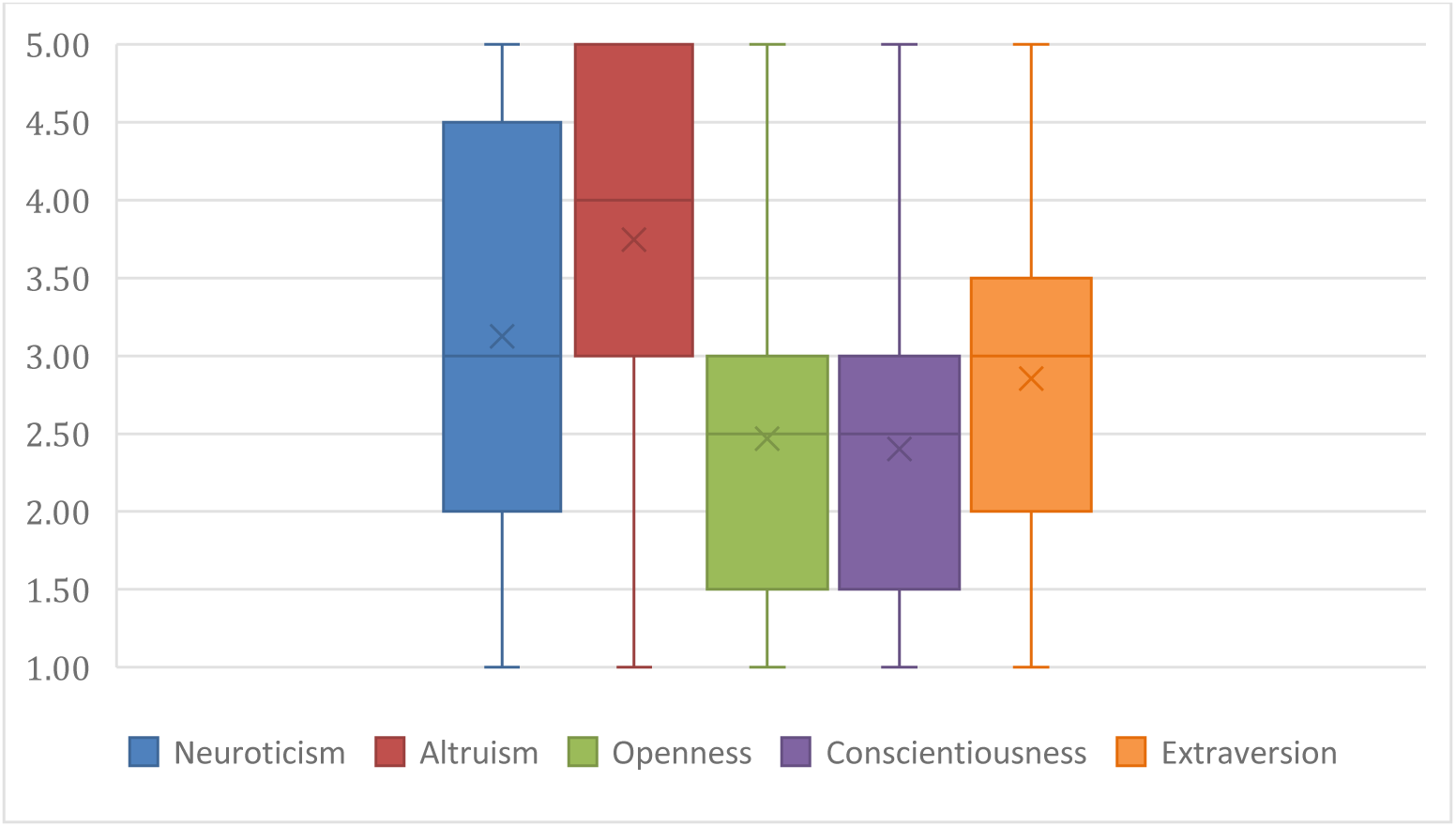
**Boxplot of the representativeness of the participants on the big-five (N=214)**

**Fig 2.**
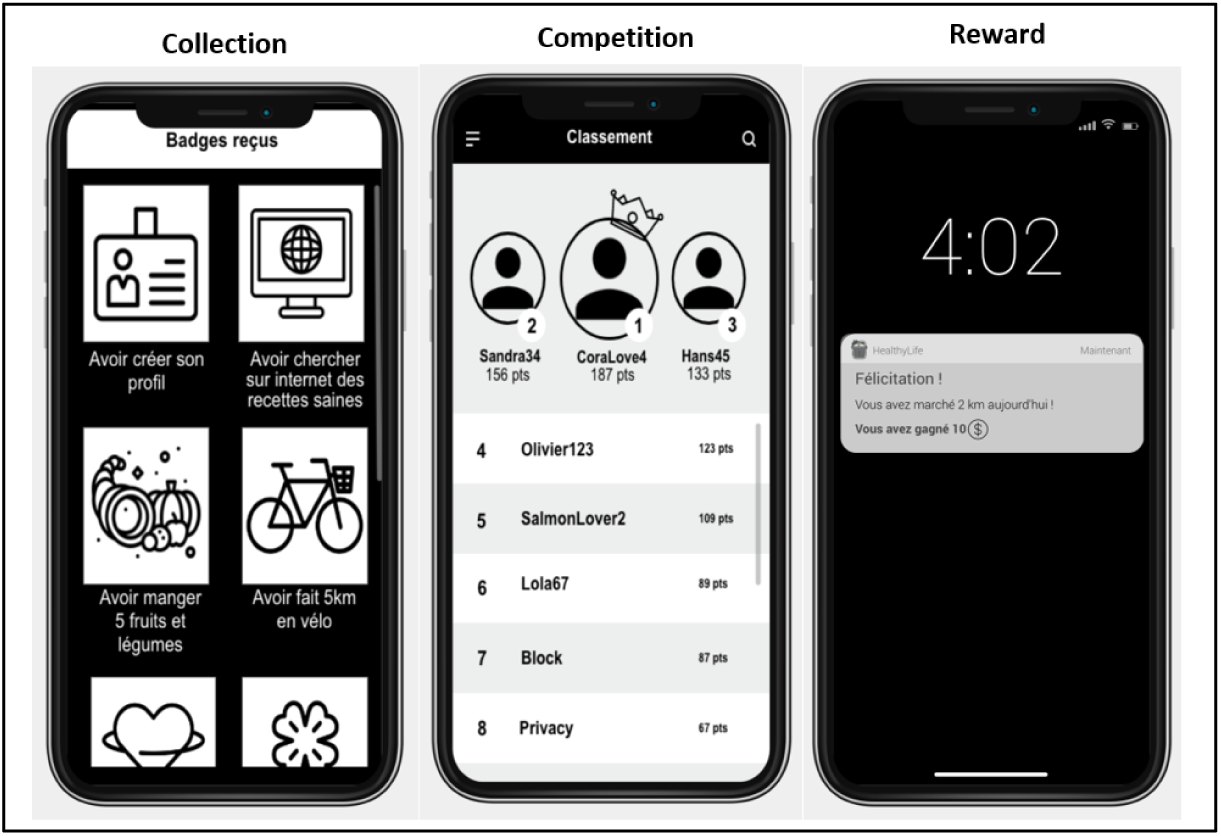
**Preferred mechanisms according to personality profiles**

### Explanation of choice

For each of the 5 selected mechanisms, participants were requested to indicate the extent to which that mechanism would motivate them to engage in healthier behaviors on a scale of 0 to 100. Subsequently, participants were required to provide a free-form textual justification for their selection of the mechanisms. If a mechanism was excluded from the initial selection, it was assigned a score of zero.

## Analysis

### Preference matrix validation

In order to perform a comparative analysis of the experimental results and the initial preference matrix, the collected data were also organized into a matrix. The values of the initial matrix (M1) represent the number of preference relations observed in the literature following the scoping review that we conducted previously [7]. We modified this matrix by assigning a value of -1 to indicate an aversion relation. A value of 0 indicates that preferences and aversions were observed in the scoping review, and a null value is assigned when no relations were identified. The second matrix (M2) contains the data collected in this study. The value of the cell represents the score of the participants with the Big Five dimension for the mechanism on the line. In each cell, we calculated a mechanism discriminative score by summing the participants’ scores, divided by the total number of participants.

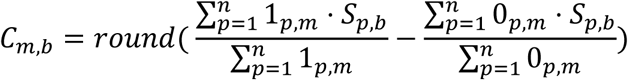

C_m,b_ represents the value in the cell corresponding to mechanism m (row) and the bigFive b (column).

1_p,m__ is an indicator function equal to 1 if participant p has selected mechanism m, and 0 otherwise.

0_p,m__ is an indicator function equal to 0 if participant p has selected mechanism m, and 1 otherwise.

S_p,b_ is the score of participant p on the big five b dimension (1-5). n is the total number of individuals.

A third matrix (M3) represents a comparison between the two matrices (M1, M2).

### Statistical analysis

A logistic regression analysis was conducted to examine the relationship between the mechanisms and the personality profile scales. The objective of this analysis was to analyze if the participants’ scores on the BFI-10-Fr significantly influences the choice of a mechanism. A logistic regression analysis was conducted for each mechanism to ascertain whether the BFI-10-Fr scores were predictive of the mechanism selection. Furthermore, a logistic ordinal regression was conducted with the motivation scores of the selected mechanisms as the dependent variables and the BFI-10-Fr scores as the predictors. The regression was performed for each mechanism motivation score, with the objective of determining whether the scores on the scales predict the mechanism score.

To prevent a type I error, the Bonferroni correction was employed for all regression analyses.

## Results

### Demographics data

A total of 214 individuals responded to the questionnaire, comprising 118 women, 89 men, 5 individuals identified as other, and 2 individuals who did not provide a response. The mean age was 29.42 years (SD = 10.41). Further details can be found in Table 4.

**Table 4.** Demographics data.

|  | **N** | **Mean** | **Standard deviation** |
| --- | --- | --- | --- |
| Age | 214 | 29.42 | 10.41 |
|  | **N** | **%** |  |
| Gender | 214 |  |  |
| Women | 89 | 41.6 |  |
| Men | 118 | 55.1 |  |
| Others | 7 | 3.3 |  |
| Education Level | 211 |  |  |
| Mandatory education | 53 | 24.8 |  |
| Bachelor's degree | 63 | 29.4 |  |
| Master's degree | 80 | 37.4 |  |
| Doctorate | 15 | 7 |  |
| Smartphone use | 214 |  |  |
| Not comfortable | 10 | 4.7 |  |
| Comfortable | 204 | 95.3 |  |
| Already used mHealth | 135 | 63.1 |  |

### Characteristics of the participants on the big-five

The scores for the Big Five dimensions are recorded on a scale of 1 to 5. The distribution of the population on the Big Five is shown in Fig 1 (the line represents the median and the cross represents the mean).

The participants of this study are mainly altruistic (M=3.71, SD= 1.31), extravert (M=2.86, SD=1.17) and neurotic (M=3.12, SD= 1.23).

### Characteristics of the participants on the selection for each mechanism

A total of four mechanisms were selected by more than half of the participants. These mechanisms included self-monitoring, progression, challenge, and quest (Table 5).

**Table 5.**
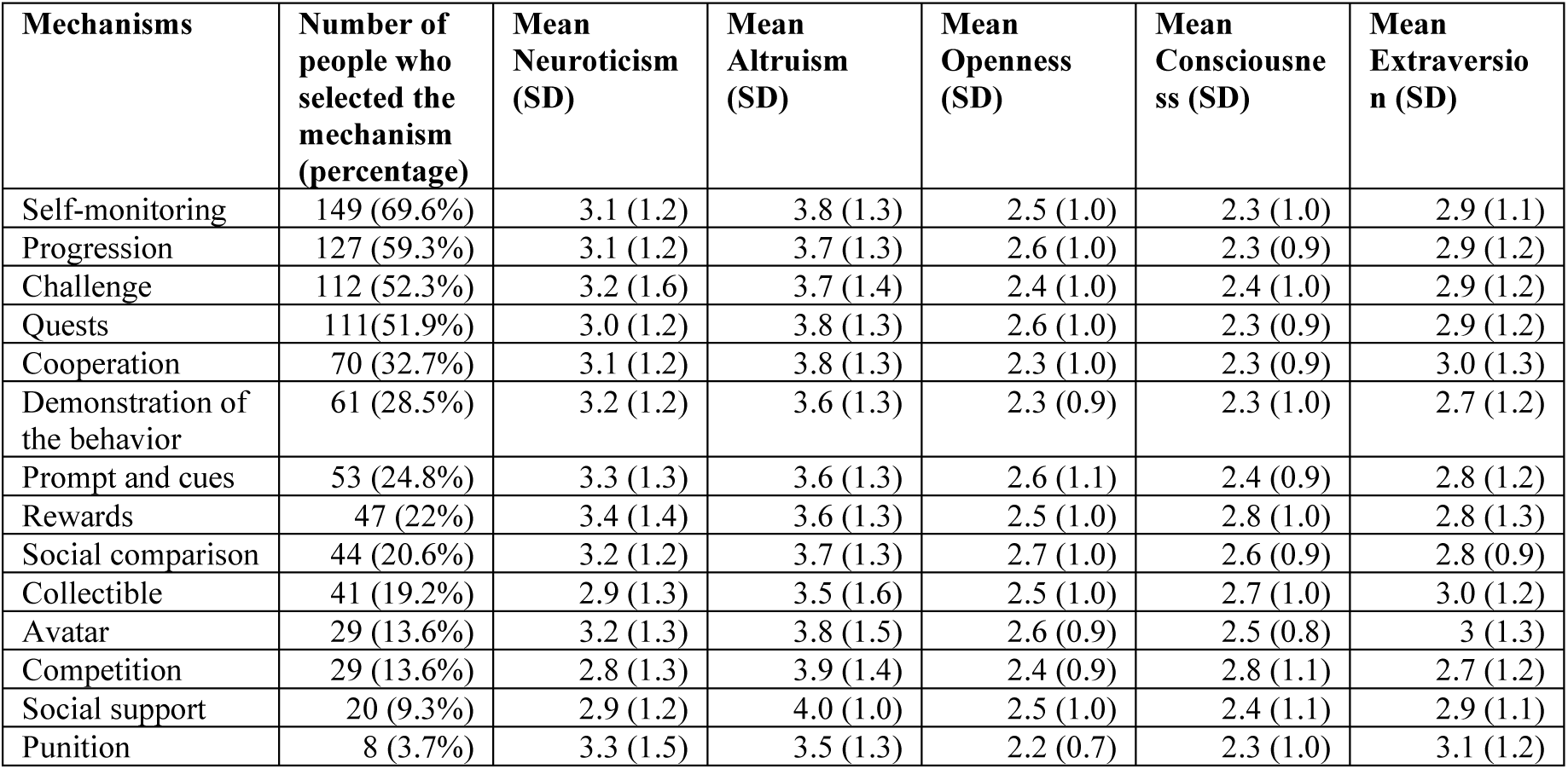
Number of people who selected the mechanism (N=214)

| Mechanisms | Number of people who selected the mechanism (percentage) | Mean Neuroticism (SD) | Mean Altruism (SD) | Mean Openness (SD) | Mean Conscientiousness (SD) | Mean Extraversion (SD) |
| --- | --- | --- | --- | --- | --- | --- |
| Self-monitoring | 149 (69.6%) | 3.1 (1.2) | 3.8 (1.3) | 2.5 (1.0) | 2.3 (1.0) | 2.9 (1.1) |
| Progression | 127 (59.3%) | 3.1 (1.2) | 3.7 (1.3) | 2.6 (1.0) | 2.3 (0.9) | 2.9 (1.2) |
| Challenge | 112 (52.3%) | 3.2 (1.6) | 3.7 (1.4) | 2.4 (1.0) | 2.4 (1.0) | 2.9 (1.2) |
| Quests | 111(51.9%) | 3.0 (1.2) | 3.8 (1.3) | 2.6 (1.0) | 2.3 (0.9) | 2.9 (1.2) |
| Cooperation | 70 (32.7%) | 3.1 (1.2) | 3.8 (1.3) | 2.3 (1.0) | 2.3 (0.9) | 3.0 (1.3) |
| Demonstration of the behavior | 61 (28.5%) | 3.2 (1.2) | 3.6 (1.3) | 2.3 (0.9) | 2.3 (1.0) | 2.7 (1.2) |
| Prompt and cues | 53 (24.8%) | 3.3 (1.3) | 3.6 (1.3) | 2.6 (1.1) | 2.4 (0.9) | 2.8 (1.2) |
| Rewards | 47 (22%) | 3.4 (1.4) | 3.6 (1.3) | 2.5 (1.0) | 2.8 (1.0) | 2.8 (1.3) |
| Social comparison | 44 (20.6%) | 3.2 (1.2) | 3.7 (1.3) | 2.7 (1.0) | 2.6 (0.9) | 2.8 (0.9) |
| Collectible | 41 (19.2%) | 2.9 (1.3) | 3.5 (1.6) | 2.5 (1.0) | 2.7 (1.0) | 3.0 (1.2) |
| Avatar | 29 (13.6%) | 3.2 (1.3) | 3.8 (1.5) | 2.6 (0.9) | 2.5 (0.8) | 3 (1.3) |
| Competition | 29 (13.6%) | 2.8 (1.3) | 3.9 (1.4) | 2.4 (0.9) | 2.8 (1.1) | 2.7 (1.2) |
| Social support | 20 (9.3%) | 2.9 (1.2) | 4.0 (1.0) | 2.5 (1.0) | 2.4 (1.1) | 2.9 (1.1) |
| Punition | 8 (3.7%) | 3.3 (1.5) | 3.5 (1.3) | 2.2 (0.7) | 2.3 (1.0) | 3.1 (1.2) |

### Comparison between this study results and the preference matrix

The initial matrix (M1) presents 60% (42/70) of positive relation, 1% (1/70) of aversion relation, 10% (7/70) of no preference relations (preference and aversion were observed) and 29% (20/70) of missing relations (see M1 on Table 6).

**Table 6.**
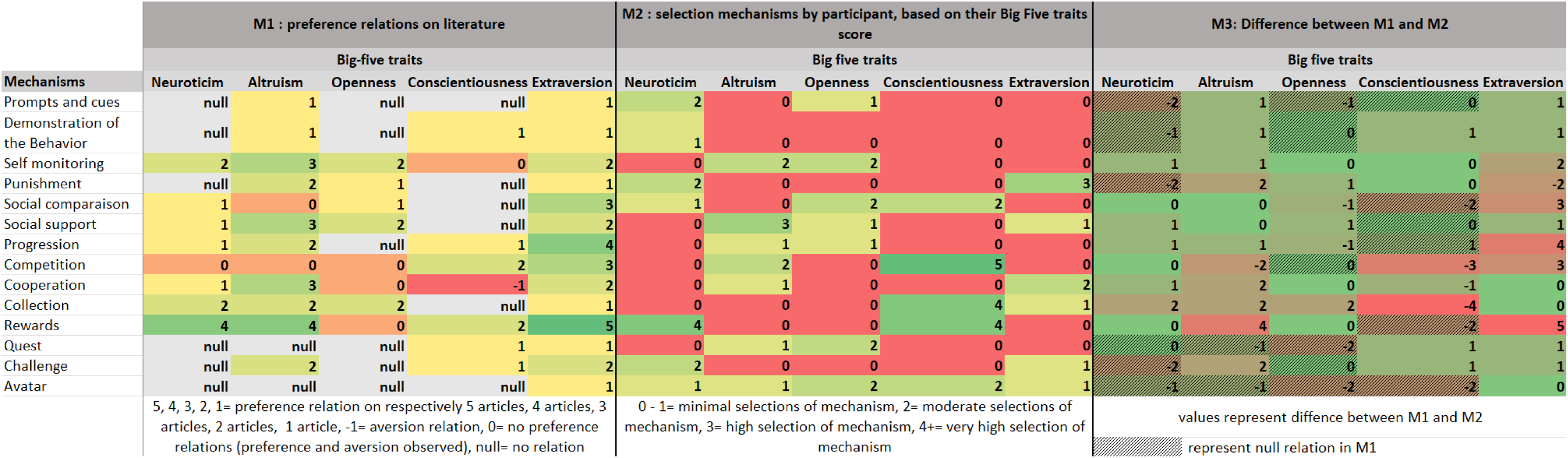
Matrices for the representation of preference relations on literature (M1), of the selection mechanisms by participants based on their Big-Five traits score (M2), d the difference between M1 and M2 (M3).

The matrix of the selection mechanisms by participant (M2) presents 5% (4/70) of very high mechanism discriminative score (<4), 3% (2/70) of high selection mechanism (3), 17% (12/70) of medium selection mechanism (2), 74% (52/70) of low selection mechanisms (0-1) (see M2 on Table 6).

The matrix M3 shows the difference between M1 and M2. The higher the value, the greater the difference between the 2 matrices. We observe 27% (19/70) similar value in the 2 matrices (no differences), 38% (27/70) minimal difference of one, 24% (17/70) moderate difference of two, and 9% (7/70) significant difference of three and more (see M3 on Table 6).

### Preferred mechanisms and big-five, statistical analysis

A logistic regression analysis was conducted for each mechanism to ascertain whether the scores on the scale were predictive of the mechanism selection. Furthermore, a logistic ordinal regression was conducted with the motivation scores of the selected mechanisms as the dependent variables and the BFI-10-Fr scores as the predictors. The logistic regression containing all predictors was statistically significant for three mechanisms.

### Collection Mechanism

The logistic regression, conducted for each mechanism to ascertain whether the scores on the big-five scale, were predictive of the mechanism selection collection, was statistically significant, 𝜒^2^(5, N=207)= 13.07, *p* <.05, indicating that the regression was able to distinguish between respondents who selected and did not select the collection mechanism. The regression as a whole explained between .06% (Cox and Snell R square) and 10% (Nagelkerke R squared) of the variance in the selection of the mechanism collection, and correctly classified 83.1% of cases. As shows in Table 7, only one of the independent variables made a unique statistically significant contribution to the model, conscientiousness with an odds ratio of 1.87. This indicated that respondents who had a high level of conscientiousness were over 1.87 times more likely to select the mechanism collection than those who have a small level of conscientiousness, controlling for all other factors in the model.

**Table 7.** Logistic Regression Predicting Likelihood of selecting the mechanism Collection.

|  | B | S.E | Wald | Df | p | Odds Ratio | 95,0% C.I for Odds Ratio |  |
| --- | --- | --- | --- | --- | --- | --- | --- | --- |
| Openness | -.019 | .193 | .010 | 1 | .920 | .981 | .672 | 1.432 |
| Conscientiousness | .625 | .198 | 9.987 | 1 | <.01 | 1.868 | 1.268 | 2.752 |
| Extraversion | .035 | .168 | .043 | 1 | .836 | 1.035 | .745 | 1.439 |
| Altruism | -.165 | .146 | 1.279 | 1 | .258 | .848 | .637 | 1.129 |
| Neuroticism | -.191 | .163 | 1.376 | 1 | .241 | .826 | .600 | 1.137 |
| Constant | -2.089 | 1.093 | 3.654 | 1 | .056 | .124 |  |  |

### Competition Mechanism

The logistic regression was statistically significant, 𝜒^2^(5, N=200) = 31.49, *p* <.01, indicating that the model was able to distinguish between respondents who selected and did not select the mechanism competition. The regression explained between 15% (Cox and Snell R square) and 33% (Nagelkerke R squared) of the variance in the selection of the mechanism collection, and correctly classified 91.5% of cases. As shows in Table 8, four of the independent variables made a unique statistically significant contribution to the regression (neuroticism, conscientiousness, extraversion, and altruism). The strongest predictors of selecting the mechanism competition were conscientiousness with an odds ratio of 3.22 and altruism with an odds ratio of 1.93. The odds ratio of .43 for neuroticism and of .52 for extraversion, indicating that the respondents with a high level of neuroticism were over .42 times less likely to select this mechanism.

**Table 8.** Logistic Regression Predicting Likelihood of selecting the mechanism Competition.

|  | B | S.E | Wald | df | p | Odds Ratio | 95,0% C.I for Odds Ratio |  |
| --- | --- | --- | --- | --- | --- | --- | --- | --- |
| Openness | -.195 | .286 | .466 | 1 | .495 | .823 | .469 | 1.441 |
| conscientiousness | 1.169 | .318 | 13.532 | 1 | <.01 | 3.220 | 1.727 | 6.003 |
| Extraversion | -.660 | .280 | 5.574 | 1 | <.05 | .517 | .299 | .894 |
| Altruism | .660 | .296 | 4.969 | 1 | <.05 | 1.934 | 1.083 | 3.454 |
| Neuroticism | -.854 | .279 | 9.364 | 1 | <.01 | .426 | .246 | .736 |
| Constant | -3.680 | 1.798 | 4.187 | 1 | <.05 | .025 |  |  |

The logistic ordinal regression conducted for each mechanism to ascertain whether the scores on the big-five scale were predictive of the motivation score for competition mechanism was statistically significant, 𝜒^2^(5, N=214)= 11.16, *p* =.05, indicating that the model was able to distinguish between participants’ scores for motivation to get fit using the competition mechanism. The model explained between 5% (Cox and Snell R square) and 8% (Nagelkerke R squared) of the variance in the selection of the mechanism competition. As shown in Table 9, two of the independent variables made a unique statistically significant contribution to the regression (neuroticism, and conscientiousness). The strongest predictor of selecting the mechanism competition was conscientiousness with an odds ratio of 1.63 and neuroticism with an odds ratio of 0.71.

**Table 9.** Logistic Regression Predicting Likelihood of score of the mechanism Competition.

|  | B | S.E | Wald | df | p | Odds Ratio | 95,0% C.I for Odds Ratio |  |
| --- | --- | --- | --- | --- | --- | --- | --- | --- |
| Openness | -.072 | .207 | .121 | 1 | .728 | .931 | -.478 | .553 |
| conscientiousness | .487 | .206 | 5.586 | 1 | **.018** | 1.628 | .083 | .892 |
| Extraversion | -.272 | .191 | 2.028 | 1 | .154 | .762 | -.647 | .102 |
| Altruism | .206 | .177 | 1.357 | 1 | .244 | 1.229 | -.141 | .553 |
| Neuroticism | -.345 | .179 | 3.705 | 1 | **.054** | .708 | -.696 | .006 |

### Rewards Mechanism

The logistic regression was statistically significant, 𝜒^2^(5, N=204)= 35.29, *p* <.001, indicating that the model was able to distinguish between respondents who selected and did not select the mechanism rewards. The regression explained between 16% (Cox and Snell R square) and 27% (Nagelkerke R squared) of the variance in the selection of the mechanism rewards, and correctly classified 81.4% of cases. As shown in Table 10, two of the independent variables made a unique statistically significant contribution to the model (neuroticism, and conscientiousness). The strongest predictors of scoring the mechanism competition were conscientiousness with an odds ratio of 2.36 and neuroticism with an odds ratio of 1.97. The odds ratio indicating that the respondents with a high level of neuroticism were over 1.97 times more likely to select this mechanism and respondents with high level of conscientiousness were over 2.36 times more likely to select this mechanism.

**Table 10.** Logistic Regression Predicting Likelihood of selecting the mechanism rewards.

|  | **B** | **S.E** | **Wald** | **df** | **p** | **Odds Ratio** | **95,0% C.I for Odds Ratio** |  |
| --- | --- | --- | --- | --- | --- | --- | --- | --- |
| Openness | .060 | .194 | .095 | 1 | .758 | 1.061 | .726 | 1.551 |
| conscientiousness | .858 | .220 | 15.262 | 1 | <.01 | 2.360 | 1.534 | 3.630 |
| Extraversion | -.220 | .187 | 1.392 | 1 | .238 | .802 | .557 | 1.157 |
| Altruism | .068 | .161 | .177 | 1 | .674 | 1.070 | .781 | 1.466 |
| Neuroticism | .679 | .190 | 12.696 | 1 | <.01 | 1.971 | 1.357 | 2.864 |
| Constant | -5.919 | 1.327 | 19.897 | 1 | <.01 | .003 |  |  |

The logistic ordinal regression was statistically significant, 𝜒^2^(5, N=214)= 14.36, *p* =.01, indicating that the model was able to distinguish between participants’ scores for motivation to get fit using the reward mechanism. The regression explains between 6% (Cox and Snell R square) and 9% (Nagelkerke R squared) of the variance in the selection of the mechanism rewards. As shows in Table 11, two of the independent variables made a unique statistically significant contribution to the regression (neuroticism, and conscientiousness). The strongest predictors of scoring the mechanism rewards was conscientiousness with an odds ratio of 1.66 and neuroticism with an odds ratio of 1.38.

**Table 11.** Logistic Regression Predicting Likelihood of the score the mechanism rewards.

|  | B | S.E | Wald | df | p | Odds Ratio | 95,0% C.I for Odds Ratio |  |
| --- | --- | --- | --- | --- | --- | --- | --- | --- |
| Openness | .025 | .170 | .023 | 1 | .881 | 1.026 | -.307 | .358 |
| conscientiousness | .504 | .176 | 8.199 | 1 | <.01 | 1.66 | .159 | .849 |
| Extraversion | -.031 | .156 | .040 | 1 | .842 | .969 | -.336 | .274 |
| Altruism | -.083 | .132 | .389 | 1 | .533 | .921 | -.342 | .177 |
| Neuroticism | .320 | .149 | 4.606 | 1 | <.05 | 1.378 | .028 | .61 |

## Discussion

The objective of this study was to validate the preference matrix obtained through a previous literature review by observing if we obtain similar relations within this study.

It was observed that four mechanisms were selected by more than half of the participants: self-monitoring (N=149), progression (N=127), challenge (N=112), and quest (N=111).

No significant relationship was identified between these mechanisms and the Big Five dimensions. It can thus be inferred that these four mechanisms are appreciated by any type of participants and should be included in each mHealth application independantly of the profile of its users.

### Comparison between M1 and M2

The comparison between matrices M1 and M2 reveals several noteworthy developments in the identification of potential relationships between mechanisms and Big Five personality traits. In M2, new possible associations emerge that were not identified in M1, particularly between neuroticism and mechanisms such as prompts and cues, demonstration of behavior, punishment, quest, challenge, and avatar. Similarly, openness is newly associated with prompts and cues, progression, quest, and challenge, while conscientiousness shows novel relations with social comparison and avatar, and a notably strong association with collection in M2. These findings suggest an evolving understanding of how specific personality traits may align with certain mechanisms. Furthermore, while M1 lacked a clear consensus regarding the relationships between mechanisms and traits such as altruism and openness, M2 suggests emerging relations— particularly between altruism and competition, and between openness and rewards. Conversely, several relationships identified in M1 do not reappear in M2. For example, extraversion was previously associated with a wide range of mechanisms in M1— including prompts and cues, demonstration of behavior, self-monitoring, punishment, social comparison, social support, progression, competition, cooperation, collection, rewards, and quest—but these associations are not observed in M2. A similar decline in associations is seen for altruism, which in M1 was linked with prompts and cues, demonstration of behavior, punishment, collection, challenge, and especially rewards. However, significant results were only found for the mechanisms collection, competition, and rewards. Three of the five significant relationships found in this study were present in the preference matrix M1, conscientiousness with competition, conscientiousness with reward and neuroticism with rewards.

Two have not a consensus on the preference matrix M1 with articles observed preference and non-preference relations between neuroticism with competition, and altruism with competition.

This study permits to enrich the preference matrix with the addition of one new significative relation between conscientiousness with collection. And to confirm when literature observe incongruent data, we clarify with our results a preference relation for neuroticism with competition, and altruism with competition.

Below we explore in more detail the three mechanisms that yielded significant regressions. Fig 3 depicts the three mechanisms, which were presented to participants in the questionnaire with a brief description.

### Collection

In our context, the term “collection” refers to a set of items or badges that are accumulated over time. The act of collecting as a mechanism that can serve as a motivational tool for stay fit [28]. In our study conscientious participants selected 1.87 times collection than those who have a small level of conscientiousness.

These findings are corroborated by the existing literature. In the preference matrix M1, the collection mechanism was preferred by all big-five traits [9,23,29–31].

### Competition

The competition mechanism can be defined as the fact that users are able to engage in a competitive process with one another in order to perform the desired behavior [22].

This mechanism is related to all Big Five traits in the preference matrix M1. But in this study, individuals exhibiting conscientious (OR= 3.22) and altruistic tendencies (OR=1.93) were more likely to identify this mechanism as a motivating factor in maintaining their health than participant who have a small level of these dimensions. In contrast, extroverts (OR=.42) and individuals with neurotic tendencies (OR=.52) exhibited a reduced propensity to select this mechanism. Furthermore, respondents exhibiting high levels of conscientiousness (OR=1.63) assigned higher ratings to this mechanism, whereas those with neurotic personalities (OR=0.71) assigned lower ratings.

These differences can be explained by a controversial preference for this mechanism. Some participants said they preferred individual mechanisms that didn’t involve other users, they involved personal, individual efforts to motivate to stay fit. The results is in line with previous research that found similar reasons against competition with their participants [10].

> *[P14] “No comparison with others because it stresses me out if I don’t win. I’m more motivated by the idea of doing myself good”.*

> *[P39] “I choose for myself, not for others, so there’s no competition or comparison with others.”*

> *[P129] “I’ve excluded those that involve other people because I prefer autonomy.” [P134] “I no longer have anything to prove to anyone but myself”.*

> *[P112] “The individuality aspect. In my opinion, when you do something, you do it for yourself and not for others”.*

> *[P140] “I think the principle of fitness should be personal and not a competition with others.”*

While other participants said they found competition more motivating:

> *[P44] “I find competition very enjoyable, it pushes me to excel.”*

> *[P176] “Comparing myself to others pushes me to do better.”*

> *[P192] “I’m very competitive and I like it when people show me that I’ve made progress. It encourages me to keep going. »*

The preference matrix (M1) further indicates that altruistic and conscientious individuals tend to prefer competition [8,10,32]. However, it also demonstrates a relation of preference with the profiles of extraversion and neuroticism [10,23,32,33], whereas our study results indicate that extraverted and neurotic individuals select this mechanism with lower frequency. However, preference matrix (M1) also indicates a non-preferred relation for individuals with neuroticism profiles. The results of our experiment provide further corroboration of this relation.

### Rewards

The reward mechanism entails the provision of virtual incentives to users for the completion of the desired action [22]. It differs from the concept of collection in that the rewards are not necessarily part of a set of collectible items.

Individuals with high levels of neuroticism (OR=1.97) and conscientiousness (OR=2.36) demonstrated a greater propensity to select the rewards mechanism.

Additionally, they assigned a higher motivation score to this mechanism (OR_Conscientiousness_=1.66; OR_Neuroticism_=1.38).

Preference matrix (M1) reveals a preference relation between these two profiles and the rewards [23,30,32–35].

### Dimensions du Big-Five

It is noteworthy that the conscientiousness trait is a significant predictor of all three mechanisms.

No significant results were obtained regarding openness of participants, despite the existing literature indicating a preference for specific mechanisms like collection and competition.

No significant preference results were obtained with the extraversion traits. Our preference matrix, however, does exhibit relations for all mechanisms. A summary of results and preference matrix is presented on table 12.

**Table 12.** Mechanisms preferred and non-preferred per big-five dimension (in bold congruent finding between our test and the literature, in italic incongruent).

| Big-five dimension | Mechanisms preferred on this study | Mechanisms non-preferred on this study | Mechanisms preferred on preference matrix (M1) | Mechanisms non-preferred on preference matrix (M1) |
| --- | --- | --- | --- | --- |
| Openness to experience | - | - | Self-monitoring, punishment, social comparison, social support, **competition**, cooperation, **collection**, **rewards**, customization. | Rewards, *competition*, cooperation |
| Agreeableness | *Competition* | - | Prompts and cues, demonstration of the behavior, self-monitoring, punishment, social comparison, social support, progression, **competition**, cooperation, collection, rewards, challenge, customization. | Social comparison, *competition* |
| Conscientiousness | **Collection ; Competition ; Rewards** | - | Demonstration of the behavior, self-monitoring, progression, **competition**, **collection**, **rewards**, quest, challenge. | Self-monitoring, cooperation, customization |
| Extraversion | - | *Competition* | Prompts and cues, demonstration of the behavior, self-monitoring, punishment, social comparison, social support, progression, *competition*, cooperation, collection, rewards, quest, challenge, avatar, customization. | customisation |
| Neuroticism | **Rewards** | *Competition* | Self-monitoring, social comparison, social support, progression, *competition*, cooperation, collection, **rewards**, customization. | **competition** |

### Limitations

The fact that participants had to select five mechanisms out of fifteen was a limitation. Some participants may have selected mechanisms they didn’t find motivating because they had to select five. Others, on the contrary, might have selected more.

Our population is not representative of the general population, due to its high inclusion of university students, a demographic that is typically considered to be relatively young (M_age_=29.42, SD_age_=10.41).

We also observe that, despite our sample size calculation, it appears that it did not allow for proper discrimination, as we obtained significant results for only 3 out of 14 mechanisms. Therefore, this calculation should be reconsidered.

It was not feasible to assess the customization mechanism, given its expansive scope and the consequent challenges in representing it in the format of a conventional mockup screen.

## Conclusion

The entire preference matrix could not be validated experimentally. Significant relations were identified for three mechanisms: collection, competition, and rewards. The preference relations are corroborated by the preference matrix M1, apart from preference relation for collection mechanism and conscientiousness and the non-preference relation for competition regarding the extraversion trait, which is significant in this study but, according to the preference matrix M1, is a preference relation. The neuroticism trait is significant for competition as non-preferred, yet the preference matrix studies demonstrated a preference and non-preference relation for this trait. Considering these findings, it can be concluded that a non-preference relation is corroborated. It would be interesting to replicate this study with a larger, less student-dominated panel to get better representation and more significant results on mechanism preference relations by Big Five traits.

## Data availability

The data sets generated during the current study are available from the corresponding author on reasonable request.

## Acknowledgment

None declared.

## Authors’ contributions

LG conceived the study with the involvement and advice of FE and GF. MP is involved with statistics. All authors were involved in writing, reading, and approving the final manuscript.

## Multimedia Appendix

S1 Appendix: Print version of the online questionnaire

